# Rapid emergence of transmissible SARS-CoV-2 variants in mild community cases

**DOI:** 10.1101/2023.02.15.23285923

**Authors:** Michael A Crone, Seran Hakki, Jie Zhou, Carolina Rosadas de Oliveira, Kieran J Madon, Aleksandra Koycheva, Anjna Badhan, Jakob Jonnerby, Joe Fenn, Rhia Kundu, Jack L Barnett, Sean Nevin, Emily Conibear, Nieves Derqui-Fernandez, Timesh D Pillay, Robert Varro, Constanta Luca, Valerie Quinn, Shazaad Ahmad, Maria Zambon, Wendy S Barclay, Jake Dunning, Paul S Freemont, Graham P Taylor, Ajit Lalvani

## Abstract

SARS-CoV-2 immune-escape variants have only been observed to arise in immunosuppressed COVID-19 cases, during prolonged viral shedding. Through daily longitudinal RT-qPCR, quantitative viral culture and sequencing, we observe for the first time the evolution of transmissible variants harbouring mutations consistent with immune-escape in mild community cases within 2 weeks of infection.

## Introduction

Mutations have been observed to develop over the course of several weeks or months in chronically infected patients in hospital with haematological malignancies and iatrogenic immunosuppression (1) and in community cases with advanced acquired immunodeficiencies (2). Our current understanding of how SARS-CoV-2 immune-escape variants arise is that the virus is not sufficiently supressed by the immune system in profoundly immunosuppressed individuals, allowing the virus to develop non-synonymous mutations through ongoing viral evolution (3) leading to immune-escape (1,2,4,5).

To determine whether such variants with infectious potential arise in mild community COVID-19 cases without advanced immunosuppression, we aimed to investigate whether infectious variants develop during infection by sequencing and culturing of SARS-CoV-2 isolates from serial nose and throat samples in a community cohort.

## The Study

Between May 2021 and October 2021, during which the Delta variant (B.1.617.2) was the UK’s dominant strain, 343 community contacts of symptomatic PCR-confirmed index cases were recruited within five days of their index case’s symptom onset to the ATACCC study as previously described (6,7). 78/343 (23%) of these delta-exposed contacts were determined to be PCR-positive for more than one timepoint. Of these, 32 cases had their viral growth phase captured (6). Viral culture plaque assays and lateral flow devices (Innova, Xiamen, PRC) were performed on thawed viral transport media containing nasopharyngeal swabs, as previously described (6).

We performed longitudinal SARS-CoV-2 whole-genome sequencing (WGS) using an amplicon-based WGS protocol in the eleven of 32 cases (Table S1) who had at least four consecutive samples with over 1,000 RNA viral copies/mL on RT-qPCR. Criteria to determine the significance of SARS-CoV-2 mutations were: (i) detection of mutation in at least 5% of sequencing reads at a particular position and (ii) mutant virus proportion altered over time and (iii) existence of published reports identifying the mutation as a candidate immune escape variant.

Of the eleven cases, nine (6 vaccinated; 3 unvaccinated) shed infectious virus for a typical duration (5 days [IQR 3-7]) (6) and no mutations meeting our criteria were detected, although one case, Case-E, developed a mutation (ORF1ab:T283I) shown to have a neutral effect (8).

Two vaccinated cases, Case-A and B showed persistence of infectious virus until the end of the sampling period (13-days and 10-days of plaque assay positivity for Case-A and B respectively) despite concurrent negative lateral flow tests during the latter stages of infection (Fig.1).

Notably, both developed multiple significant mutations. The mutations detected were in ORF7a (two large deletions, DF54-Q62insL and DA55-C67), a gene which blocks the incorporation of the antiviral host protein SERINC5 in virions (6), and the S gene (two nonsynonymous mutations, D253G and S255F), where mutations are known to cause immune-escape (9).

**Fig. 1.**
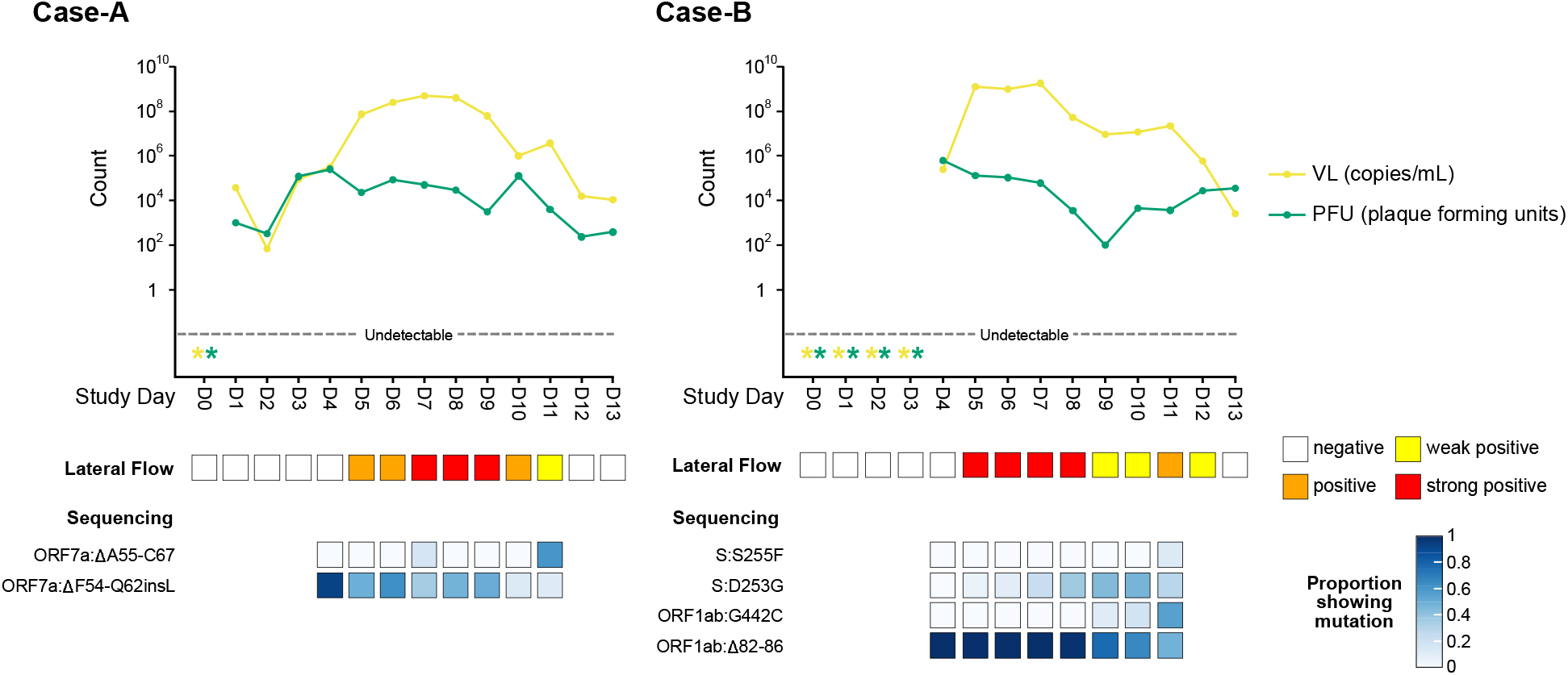
SARS-CoV-2 viral dynamics captured through daily sampling for Case-A and Case-B and a sequencing summary for nucleotide polymorphisms and indels in coding genes. Diagrams show the change in RT-qPCR viral load (VL) (yellow lines) and number of plaque forming units (PFU) obtained using a plaque assay over the course of infection (green lines). Undetectable viral load and PFU measurements are indicated by a star (*). Lateral flow results show the intensity of bands present on testing over the course of infection. Sequencing summarises results for any single nucleotide polymorphisms and indels in coding genes that met all three of our significance criteria (the mutation needed to be found in at least 5% of the sequencing reads at a particular position, the mutant virus proportion needed to increase or decrease over time, and, the mutation had to be within genes where its effect had been previously described or where mutations could play a role in immune-escape based on gene function). All viral mutations for Case-A and Case-B can be visualised in Supplementary Fig.S1-2, respectively.

Remarkably, Case-A and B both had type II diabetes mellitus and were overweight (BMI of 27-28), whereas none of the other 32 cases with daily RT-qPCR and viral culture had type II diabetes. Both Case-A and B had good glycaemic control, maintaining HbA1c levels below 48 mmol/mL for the 4 years pre-infection (Fig.S3). Neither case had a history of immunosuppression or recurrent infections. Both recovered spontaneously from mild COVID-19 and neither required hospital care.

## Conclusions

The emergence of viral mutations in severely immunodeficient patients with very prolonged viral shedding is believed to be a significant source of highly infectious SARS-CoV-2 variants. We observed that community contacts can rapidly develop viable, culturable mutations with potential for immune escape during a normal course of infection within two weeks of disease onset. This demonstrates, to our knowledge for the first time that it is feasible for immune-escape mutations to occur in community cases without advanced immunosuppression.

The only cases with diabetes in our cohort of mild self-resolving COVID-19 cases were the two in whom mutations arose and both had a longer-than-average duration of infectious viral shedding. Case-A developed mutations in ORF7a. Whilst the function of ORF7a and its potential role in immune-escape is unclear, it has recently been shown that ORF7a inhibits the incorporation of the antiviral host factor Serine Incorporator 5 (SERINC5) into budding virions (10). Deletion mutants were shown to preserve this inhibitory function (10); however, any further effects of these deletions on host antiviral inhibition have not been investigated. Although mutations within the spike protein receptor binding domain are considered to play the most important role in immune-escape (11), the two mutations observed in close genomic proximity in Case-B (D253G and S255F) occur in the Spike N-terminal domain. Mutations in this region impact binding of potent neutralising monoclonal antibodies against spike protein (9,12) with S255F detected in chronically infected cases (13) and D253G reported in a variant of interest (14).

The development of these mutations whilst the patients were still infectious (determined by continued presence of culturable virus) demonstrates that these mutations could potentially be transmitted. Case-B showed increases in viral loads at around the time when the mutations were first detectable, with increases in PFU on plaque assay and viral RNA load on RT-qPCR, that could not be accounted for by sampling variability. Viral rebound, where viral load levels increase from borderline positive or negative values, has been shown to correlate with the emergence of immune-escape (1) and this temporal correlation suggests that the development of immune-escape mutations was driving prolonged viral shedding, rather than the immune-escape variants developing because of prolonged viral shedding.

Interestingly, our results also suggest that both cases were initially infected by several haplotypes of the same quasispecies. Since it is unlikely that the deletions detected early in infection had reverted to their ancestral state, their relative decline in frequency during the course of infection is better explained by an increase in frequency under immune pressures of additional original haplotypes lacking these deletions. This interpretation is consistent with recent estimates of transmission bottleneck indicating that between one and eight viruses can be transmitted (15) with immunocompromised recipients possibly representing the higher end of that spectrum.

Our study has several limitations. Blood was not collected from the study participants; therefore plaque reduction neutralization tests (PRNT) using serial serum samples could not be carried out to specifically demonstrate the development of immune-escape properties with the accumulation of mutations. And since, for case B, we do not know if antibody responses to the Spike N-terminal domain were present prior to infection it is unclear if vaccination placed additional selection pressure on the development of the mutations.

Whilst our cohort size was small, our finding that mild, self-resolving community cases can rapidly give rise to potential immune-escape variants while still shedding infectious virus suggests a new, community-based source for the emergence and spread of SARS-CoV-2 variants. This may have impacted the course of the pandemic and the emergence of successive highly infectious variants in ways hitherto unappreciated. Whilst these variants arose from mild community cases during the delta wave, there is no *a priori* reason to believe that this would not occur in other dominating variants in circulation, such as Omicron. Given the high prevalence of type II diabetes mellitus, future surveillance with WGS in community cases should now determine whether diabetes is a significant risk factor for the emergence of immune-escape variants.

## Data Availability

All data produced in the present study are available upon reasonable request to the authors. Sequencing data will be submitted to a repository.

## Acknowledgements

This work is supported by the NIHR (NIHR200927), a Department of Health and Social Care COVID-19 Fighting Fund award, and the NIHR Health Protection Research Units (HPRUs) in respiratory infections and in modelling and health economics. The authors would like to acknowledge funding provided through PROTECT COVID-19 National Core Study and Genotype-to-Phenotype UK National Virology Consortium (G2P-UK) funded by the Medical Research Council (MRC; MR/W005611/1), which also partially funded this work. PSF and MAC are supported by the UK Dementia Research Institute which receives its funding from UK DRI Ltd, funded by the UK Medical Research Council, Alzheimer’s Society and Alzheimer’s Research UK. JD is supported by the NIHR HPRU in emerging and zoonotic infections. GPT is supported by the Imperial NIHR Biomedical Research Centre. We thank all the participants who were involved in the study, UK Health Security Agency staff for facilitating recruitment into the study, the staff of the Virus Reference Department for doing the PCR and sequencing assays, and the Immunisations Department for assisting with analysis of vaccination data. We would like to thank all the participants who were involved in the study. We thank for Alexandra Kondratiuk, Anjeli Ketkar, Berenice Di Biase, Charlotte Williams, Chitra Tejpal, Eimear McDermott, Giulia Miserocchi, Jada Samuel, Janakan Sam Narean, Jessica Russell, Harriet Catchpole, Holly Grey, Koji Nixon, Kristel Timcang, Mica Tolosa-Wright, Michael Whitfield, Mohammed Essoussi, Niamh Nichols, Samuel Bremang, Sarah Hammett, Samuel Evetts and Tamara Hopewell for conducting participant recruitment, data entry and or quality control.

## References

1. Harari S, Tahor M, Rutsinsky N, Meijer S, Miller D, Henig O, et al. Drivers of adaptive evolution during chronic SARS-CoV-2 infections. Nat Med. 2022;1–8.

2. Cele S, Karim F, Lustig G, San JE, Hermanus T, Tegally H, et al. SARS-CoV-2 prolonged infection during advanced HIV disease evolves extensive immune escape. Cell Host Microbe. 2022;30(2):154–162.e5.

3. Wilkinson SAJ, Richter A, Casey A, Osman H, Mirza JD, Stockton J, et al. Recurrent SARS-CoV-2 Mutations in Immunodeficient Patients. Virus Evol. 2022;8(2):veac050..

4. Kemp SA, Collier DA, Datir RP, Ferreira IATM, Gayed S, Jahun A, et al. SARS-CoV-2 evolution during treatment of chronic infection. Nature. 2021;592(7853):277–82.

5. Corey L, Beyrer C, Cohen MS, Michael NL, Bedford T, Rolland M. SARS-CoV-2 Variants in Patients with Immunosuppression. New Engl J Med. 2021;385(6):562–6.

6. Hakki S, Zhou J, Jonnerby J, Singanayagam A, Barnett JL, Madon KJ, et al. Onset and window of SARS-CoV-2 infectiousness and temporal correlation with symptom onset: a prospective, longitudinal, community cohort study. Lancet Respir Medicine. 2022;

7. Singanayagam A, Hakki S, Dunning J, Madon KJ, Crone MA, Koycheva A, et al. Community transmission and viral load kinetics of the SARS-CoV-2 delta (B.1.617.2) variant in vaccinated and unvaccinated individuals in the UK: a prospective, longitudinal, cohort study. Lancet Infect Dis. 2021;

8. Das JK, Sengupta A, Choudhury PP, Roy S. Characterizing genomic variants and mutations in SARS-CoV-2 proteins from Indian isolates. Gene Reports. 2021;25:101044.

9. McCallum M, Marco AD, Lempp FA, Tortorici MA, Pinto D, Walls AC, et al. N-terminal domain antigenic mapping reveals a site of vulnerability for SARS-CoV-2. Cell. 2021;184(9):2332–2347.e16.

10. Timilsina U, Umthong S, Ivey EB, Waxman B, Stavrou S. SARS-CoV-2 ORF7a potently inhibits the antiviral effect of the host factor SERINC5. Nat Commun. 2022;13(1):2935.

11. Harvey WT, Carabelli AM, Jackson B, Gupta RK, Thomson EC, Harrison EM, et al. SARS-CoV-2 variants, spike mutations and immune escape. Nat Rev Microbiol. 2021;19(7):409–24.

12. Mathema B, Chen L, Wang P, Cunningham MH, Mediavilla JR, Chow KF, et al. Genomic Epidemiology and Serology Associated with a SARS-CoV-2 R.1 Variant Outbreak in New Jersey. Mbio. 2022;e02141–22.

13. Williamson MK, Hamilton F, Hutchings S, Pymont HM, Hackett M, Arnold D, et al. Chronic SARS-CoV-2 infection and viral evolution in a hypogammaglobulinaemic individual. Medrxiv. 2021;2021.05.31.21257591.

14. West AP, Wertheim JO, Wang JC, Vasylyeva TI, Havens JL, Chowdhury MA, et al. Detection and characterization of the SARS-CoV-2 lineage B.1.526 in New York. Nat Commun. 2021;12(1):4886.

15. Lythgoe KA, Hall M, Ferretti L, Cesare M de, MacIntyre-Cockett G, Trebes A, et al. SARS-CoV-2 within-host diversity and transmission. Science. 2021;372(6539):eabg0821.

