## Supplementary Materials for "Rapid emergence of transmissible SARS-CoV-2 variants in mild community cases"

### Supplementary Information: Rapid emergence of SARS-CoV-2 immune escape variants in mild community cases

| Case | Vaccination Status | Prolonged or Normal Duration Shedding | Day | WHO Variant (Pango Lineage[1]) | Viral Load (Ct) | Viral Load (copies/mL) | Average Read Depth (x) |
|---|---|---|---|---|---|---|---|
| A | Vaccinated | Prolonged | 4 | Delta (AY.4.3) | 26.91 | 316943 | 9200.06 |
| | | | 5 | | 19.18 | 73869974 | 10456.17 |
| | | | 6 | | 17.47 | 246714125 | 8228.39 |
| | | | 7 | | 16.50 | 488965696 | 6244.98 |
| | | | 8 | | 16.78 | 401348899 | 6132.5 |
| | | | 9 | | 19.43 | 61929819 | 15836.73 |
| | | | 10 | | 25.28 | 1000473 | 6364.92 |
| | | | 11 | | 23.44 | 3662241 | 10416.98 |
| B | Vaccinated | Prolonged | 4 | Delta (AY.4.3) | 27.26 | 247620 | 3920.69 |
| | | | 5 | | 15.17 | 1249168110 | 5890.48 |
| | | | 6 | | 15.52 | 975945340 | 7903.88 |
| | | | 7 | | 14.66 | 1789863416 | 6625.62 |
| | | | 8 | | 19.67 | 52287081 | 6869.57 |
| | | | 9 | | 22.14 | 9160120 | 8051.23 |
| | | | 10 | | 21.77 | 11891100 | 7490.03 |
| | | | 11 | | 20.89 | 22117798 | 6830.64 |
| C | Vaccinated | Normal | 1 | Delta (AY.4.3) | 24.27 | 2039580 | 12704.37 |
| | | | 2 | | 21.43 | 15113140 | 9060.85 |
| | | | 3 | | 17.65 | 217302616 | 12872.44 |
| | | | 4 | | 19.16 | 74919246 | 9727.59 |
| | | | 5 | | 20.65 | 26196743 | 12004.11 |
| | | | 6 | | 22.27 | 8357680 | 15805.36 |
| D | Vaccinated | Normal | 9 | Delta (AY.4.3) | 16.72 | 418695631 | 7126.48 |
| | | | 10 | | 18.42 | 126251076 | 8018.33 |
| | | | 11 | | 20.34 | 32598062 | 9098.12 |
| | | | 12 | | 25.00 | 1218882 | 8772.49 |
| | | | 13 | | 24.85 | 1354885 | 11573.85 |
| E | Vaccinated | Normal | 7 | Delta (AY.4.3) | 23.56 | 3365070 | 9679.03 |
| | | | 8 | | 18.88 | 91274553 | 8117.68 |
| | | | 10 | | 20.23 | 35227495 | 5883.66 |
| | | | 11 | | 22.92 | 5284557 | 12871.5 |
| F | Vaccinated | Normal | 2 | Delta (AY.4) | 27.28 | 244152 | 6865.49 |
| | | | 3 | | 19.68 | 51919640 | 8821.04 |
| | | | 4 | | 15.62 | 909490651 | 6520.93 |
| | | | 5 | | 25.87 | 659940 | 15363.19 |
| | | | 6 | | 26.64 | 383420 | 6182.04 |
| | | | 7 | | 22.44 | 7413434 | 11428.03 |
| | | | 8 | | 25.31 | 979529 | 9636.06 |
| | | | 9 | | 27.3 | 240733 | 7385.79 |
| G | Vaccinated | Normal | 3 | Delta | 22.61 | 6575868 | 14994.35 |

| | | | | | | | |
|---|---|---|---|---|---|---|---|
| | | | 4 | (AY.4) | 15.89 | 751804030 | 4308.85 |
| | | | 5 | | 18.66 | 106593243 | 11375.29 |
| | | | 6 | | 20.48 | 29533412 | 16677.53 |
| | | | 7 | | 22.54 | 6908634 | 9283.31 |
| | | | 8 | | 25.81 | 688463 | 11418.13 |
| | | | 9 | | 27.42 | 221198 | 6602.77 |
| H | Vaccinated | Normal | 3 | Delta (AY.4) | 25.3 | 986461 | 14248.11 |
| | | | 4 | | 18.95 | 86878164 | 11918.15 |
| | | | 5 | | 17.76 | 201082823 | 12171.77 |
| | | | 6 | | 22.77 | 5874210 | 10239.61 |
| | | | 8 | | 26.9 | 319186 | 13084.34 |
| I | Unvaccinated | Normal | 1 | Delta (AY.4.3) | 25.83 | 678821 | 7099.65 |
| | | | 2 | | 17.27 | 284085145 | 9629.45 |
| | | | 3 | | 18.95 | 86878164 | 9003.9 |
| | | | 4 | | 22.51 | 7056354 | 9040.96 |
| | | | 5 | | 23.21 | 4307146 | 9905.06 |
| J | Unvaccinated | Normal | 2 | Delta (AY.98) | 23.96 | 2537962 | 14135.46 |
| | | | 3 | | 19.4 | 63253999 | 8782.97 |
| | | | 4 | | 18.26 | 141331411 | 6926.29 |
| | | | 5 | | 16.86 | 379332691 | 6276.95 |
| | | | 6 | | 17.6 | 225101600 | 11476.74 |
| | | | 7 | | 22.56 | 6811876 | 13999.46 |
| | | | 8 | | 24.36 | 1914151 | 12122.9 |
| | | | 9 | | 24.95 | 1262628 | 14620.81 |
| K | Unvaccinated | Normal | 1 | Delta (AY.4.3) | 20.15 | 37272075 | 10092.95 |
| | | | 2 | | 17.11 | 318018316 | 5217.45 |
| | | | 3 | | 17.75 | 202505908 | 9860.47 |
| | | | 4 | | 16.35 | 543524687 | 4738.62 |
| | | | 6 | | 18.6 | 111200318 | 2188.16 |
| | | | 7 | | 17.02 | 338857157 | 6267.31 |
| | | | 8 | | 21.91 | 10773179 | 4518.65 |
| | | | 9 | | 25.35 | 952284 | 7893.61 |

**Table S1. Case metadata, viral load and sequencing coverage for all samples included in the study.**

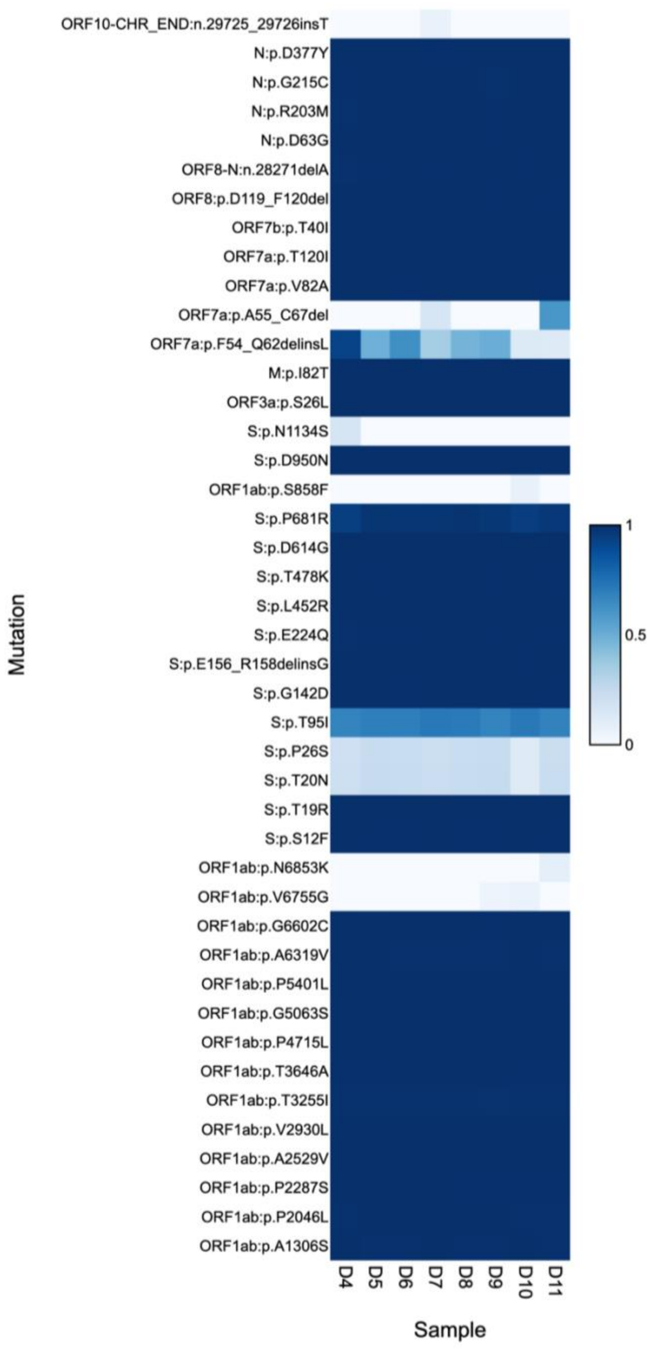

**Figure S1. Raw sequence variants called using EasySeq pipeline for Case-A.** Mutations are shown with the proportion for each from 0 (0%) to 1 (100%). Mutations are labelled in the format: {gene/region}:{protein/nucleotide}.{reference allele}{position}{alternative allele}. Partial reversion to wildtype of S:p.T95I has been previously described and is due to an amplicon dropout [2]. S:p.P26S and S:p.T20N were excluded from further analysis because they are sequencing artifacts found in all samples analysed, but were included here for completeness. None of these mutations met all three of our criteria for significance.

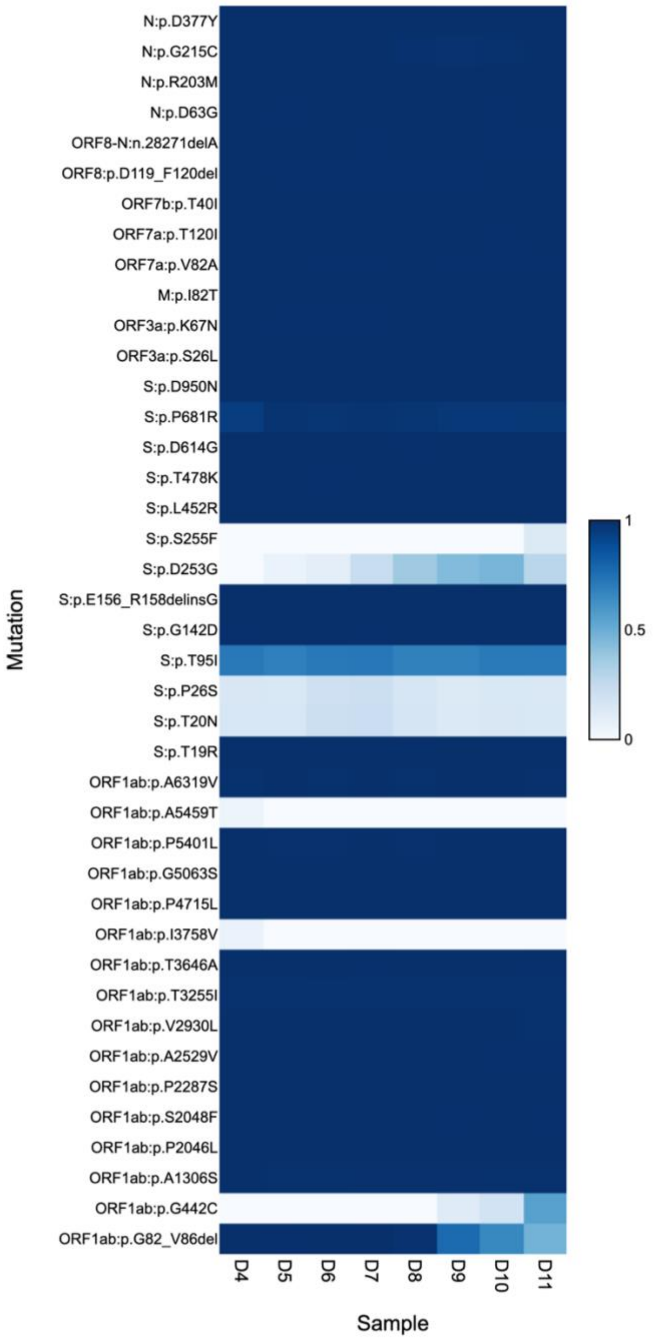

**Figure S2. Raw sequence variants called using EasySeq pipeline for Case-B.** Mutations are shown with the proportion for each from 0 (0%) to 1 (100%). Mutations are labelled in the format: {gene/region}:{**p**rotein/**n**ucleotide}.{reference allele}{position}{alternative allele}. Partial reversion to wildtype of S:p.T95I has been previously described and is due to an amplicon dropout [2]. S:p.P26S and S:p.T20N were excluded from further analysis because they are sequencing artifacts found in all samples analysed, but were included here for completeness. None of these mutations met all three of our criteria for significance.

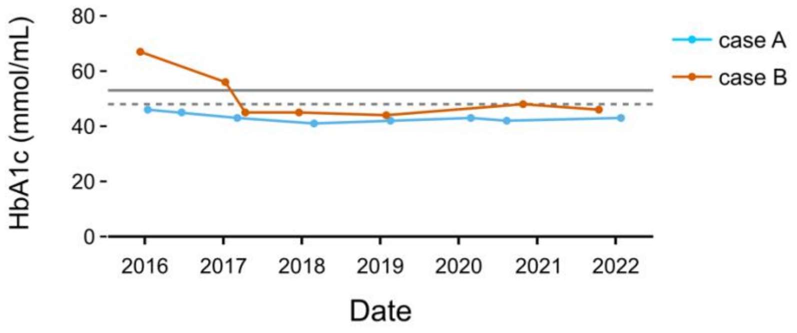

**Figure S3. Serial HbA1c measurements showing glycaemic control for both Case-A and Case-B.** The dashed grey line (48 mmol/mL) represents the NICE recommended level for those managed on diet, lifestyle and at most a single drug not associated with hypoglycaemia. The solid gray line (52 mmol/mL) is the NICE recommended level for adults on a drug associated with hypoglycaemia.

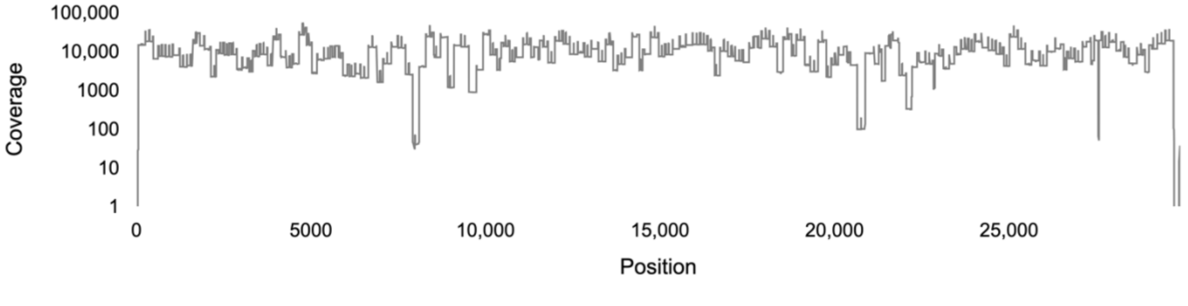

**Figure S4. Exemplar of the average sequencing read depth (shown here is Case-A Day 5) using 1 bp windows across the SARS-CoV-2 genome for the Nimagen EasySeq™ RT-PCR SARS CoV-2 (novel coronavirus) Whole Genome Sequencing kit v4.**

#### Supplementary Methods

##### RT-qPCR quantification

Automated RNA extraction was performed using a CyBio FeliX (Analytik Jena) and innuPREP Virus TS RNA Kit 2.0 (Analytik Jena) according to the manufacturer's instructions, with a sample volume of 200 µl, without carrier RNA and with an elution volume of 50 µl. RT-qPCR was repeated using an in-house protocol [3]. Samples with RT-qPCR Ct values below 28 (146,941 RNA copies per mL) were included for longitudinal sequencing (details available in Table S1).

##### WGS and data curation

cDNA synthesis was then performed using the LunaScript RT SuperMix Kit (NEB) from the RNA extracted for RT-qPCR according to the manufacturer's instructions with a total reaction volume of 20 µl and extracted sample volume of 5 µl. Sequencing libraries were generated using the EasySeq™ RT-PCR SARS CoV-2 (novel coronavirus) Whole Genome Sequencing kit v4 (Nimagen) according to the manufacturer's instructions. Samples were then pooled and purified with AMPure XP (Beckman Coulter) magnetic beads. Suitable quality of libraries was confirmed using a Tapestation (Agilent) and concentrations were measured using the Qubit 1x dsDNA High Sensitivity Assay Kit (Thermofisher Scientific) and Qubit 4 Fluorometer (ThermoFisher Scientific). The final pool was run on a NextSeq 2000 (Illumina) with a total of 322 cycles (151 bp paired reads and 10 bp indices).

Generated fastq files were processed using the EasySeq variant pipeline (v0.9) [4]. EasySeq is a Nextflow [5] pipeline that uses fastp[6], BWA MEM[7], SAMtools[8], BCFtools[8], LoFreq[9], mosdepth[10], BEDtools[11], SnpEff[12] and MultiQC [13]to QC, trim and align the reads (using reference sequence NC_045512.2) and then generate a consensus sequence and variant report before assigning a PANGO lineage [1] using pangolin (v3.1.20, lineages version 2022-02-28) [14]. Samples that failed extraction or showed any spillover during extraction were removed from further analysis. Metadata QC information from all processed samples is available in Table S1. Standard parameters were used for the pipeline, other than call_threshold which was set to 0.05 (variants were called if at least 5% of reads had the mutation present). The kit that we utilised yielded even coverage across most of the genome and an example of SARS-CoV-2 genome coverage is shown in Figure S4.

##### Plaque Assays

Briefly, thawed viral transport media (VTM) from PCR-confirmed SARS-CoV-2-positive samples were used to infect VeroE6 cell monolayers grown in 12-well plates expressing human angiotensin-converting enzyme 2 (ACE2) and transmembrane protease serine 2 precursor (TMPRSS2) (provided by MRC-University of Glasgow Centre for Virus Research (CVR), Glasgow [15]) for 1 hour. Post-infection, the inoculum was removed and cell monolayers were overlaid with a methylcellulose overlay for 72 hours before being fixed at room temperature for 20 minutes. Wells were stained for 1 hour with 1 mL of 0.05% crystal violet in 20% methanol and plaques were counted at the dilution in which there were 5-50 plaque forming units (PFUs). The limit of detection of the assay was less than 10 plaque forming units.

#### Supplementary References

1. Rambaut A, Holmes EC, O'Toole Á, et al. A dynamic nomenclature proposal for SARS-CoV-2 lineages to assist genomic epidemiology. Nat Microbiol 2020; 5:1403–1407.

2. Sanderson T, Barrett JC. Variation at Spike position 142 in SARS-CoV-2 Delta genomes is a technical artifact caused by dropout of a sequencing amplicon. Wellcome Open Res 2021; 6:305.

3. Rowan AG, May P, Badhan A, et al. Optimized protocol for a quantitative SARS-CoV-2 duplex RT-qPCR assay with internal human sample sufficiency control. J Virol Methods 2021; 294:114174.

4. Coolen JPM, Wolters F, Tostmann A, et al. SARS-CoV-2 whole-genome sequencing using reverse complement PCR: For easy, fast and accurate outbreak and variant analysis. J Clin Virol 2021; 144:104993.

5. Tommaso PD, Chatzou M, Floden EW, Barja PP, Palumbo E, Notredame C. Nextflow enables reproducible computational workflows. Nat Biotechnol 2017; 35:316–319.

6. Chen S, Zhou Y, Chen Y, Gu J. fastp: an ultra-fast all-in-one FASTQ preprocessor. Bioinformatics 2018; 34:i884–i890.

7. Li H. Aligning sequence reads, clone sequences and assembly contigs with BWA-MEM. Arxiv 2013;

8. Danecek P, Bonfield JK, Liddle J, et al. Twelve years of SAMtools and BCFtools. Gigascience 2021; 10:giab008.

9. Wilm A, Aw PPK, Bertrand D, et al. LoFreq: a sequence-quality aware, ultra-sensitive variant caller for uncovering cell-population heterogeneity from high-throughput sequencing datasets. Nucleic Acids Res 2012; 40:11189–11201.

10. Pedersen BS, Quinlan AR. Mosdepth: quick coverage calculation for genomes and exomes. Bioinformatics 2018; 34:867–868.

11. Quinlan AR, Hall IM. BEDTools: a flexible suite of utilities for comparing genomic features. Bioinformatics 2010; 26:841–842.

12. Cingolani P, Platts A, Wang LL, et al. A program for annotating and predicting the effects of single nucleotide polymorphisms, SnpEff. Fly 2012; 6:80–92.

13. Ewels P, Magnusson M, Lundin S, Käller M. MultiQC: summarize analysis results for multiple tools and samples in a single report. Bioinformatics 2016; 32:3047–3048.

14. O'Toole Á, Scher E, Underwood A, et al. Assignment of epidemiological lineages in an emerging pandemic using the pangolin tool. Virus Evol 2021; 7:veab064.

15. Rihn SJ, Merits A, Bakshi S, et al. A plasmid DNA-launched SARS-CoV-2 reverse genetics system and coronavirus toolkit for COVID-19 research. Plos Biol 2021; 19:e3001091.
